## Appendix for "Meningeal contrast enhancement in multiple sclerosis: assessment of field strength, acquisition delay, and clinical relevance"

*MRI Sequence Parameters*

7T FLAIR: TR = 8,000 ms, TE = 300 ms, TI = 2,200 ms, flip angle = 70-degrees, resolution = 0.500 x 0.488 x 0.488 mm3, SENSE acceleration factor = 2.5 x 3.0, and acquisition time = 9 minutes 4 seconds.

7T MP2RAGE: MP2RAGE TR = 8,250 ms, TR = 6.9 ms, TE= 1.97 ms, inversion times = 1,000/3,300 ms, flip angles = 5/5 degrees, Turbo factor = 252, resolution = 0.700 x 0.688 x 0.688 mm3, SENSE acceleration factor = 2 x 2, and acquisition time = 9 minutes 36 seconds.

3T FLAIR: TR = 5000 ms, TE = 390 ms, TI = 1800 ms, resolution = 0.5 x 0.5 x 1.0 mm3, GRAPPA = 2, acquisition time = 7 minutes, 27 seconds.

3T MP2RAGE: TR = 5000 ms, TE = 2.98 ms, Inversion times = 700/2500 ms, flip angles = 4/5 degrees, resolution = 1.0 x 1.0 x 1.0 mm3, GRAPPA = 3, acquisition time = 8 minutes 22 seconds.

*Supplemental Tables*

Table S1: Meningeal enhancement subtypes and total enhancement.

|  | | MS | | | HC | | |
| --- | --- | --- | --- | --- | --- | --- | --- |
|  |  | Volume (mm^3^) | Count | ME Present (%) | Volume  (mm^3^) | Count | ME Present (%) |
| Nodular | Gd+ Delayed 3T FLAIR | 2.2 (11.3)  0^b^ [0-86] | 0.1 (0.4)  0^b,c^ [0-2] | 6^b,c^ (10) | 1.6 (5.4)  0 [0-22] | 0.1 (0.3)  0 [0-1] | 2 (12.5) |
|  | Gd+ Early 7T FLAIR | 5.73 (12.4)  0^b^ [0-76.9] | 0.4 (0.7)  0^b^ [0-3] | 31^b^ (32.6) | 2.1 (7.2)  0 [0-29.5] | 0.1 (0.3)  0 [0-1] | 2 (11.8) |
|  | Gd+ Delayed 7T FLAIR | 3.3 (7.9)  0 [0-45.8] | 0.5 (1)  0^c^ [0-5] | 28^c^ (29.5) | 3.1 (7.7)  0 [0-29.9] | 0.4 (1)  0 [0-4] | 4 (23.5) |
| Spread / Fill | Gd+ Delayed 3T FLAIR | 7.5 (18.6)  0^c^ [0-95] | 0.3^c^ (0.7)  0 [0-3] | 14^c^ (23.3) | 23.9 (60.2)  0 [0-218] | 0.4 (0.6)  0 [0-2] | 5 (31.3) |
|  | Gd+ Early 7T FLAIR | 119.3 (888.6)  0^d^ [0-8623] | 0.5 (1.2)  0^d^ [0-9] | 26^d^ (27.4) | 26.3 (50.2)  0  [0-137.9] | 0.4 (0.7)  0 [0-2] | 4 (23.5) |
|  | Gd+ Delayed 7T FLAIR | 69.9 (390.5)  0^c,d^ [0-3780.7] | 0.9 (1.6)  0^c,d^ [0-10] | 42^c,d^ (44.2) | 40.8 (106.9)  0 [0-420.2] | 0.6 (1.1)  0 [0-4] | 5 (29.4) |
| Paravascular | Gd+ Delayed 3T FLAIR | 419.9 (98414)  38.4^b,c^ [0-6505] | 2.2 (3.6)  1^b,c^ [0-19] | 34^b,c^ (56.7) | 323.1 (725.5)  0^c^ [0-2837] | 1.8 (2.9)  0^c^ [0-10] | 7 (43.8) |
|  | Gd+ Early 7T FLAIR | 643.2 (999.3)  203.4^b,d^ [0-4955.1] | 3.4 (3.7)  2^a,b,d^ [0-18] | 72^b,d^ (75.8) | 143.9 (213.4)  40.7^a^ [0-728.5] | 1.3 (1.7)  1^a,d^ [0-6] | 9 (52.9) |
|  | Gd+ Delayed 7T FLAIR | 1185.7 (2058.9)  599.2^c,d^ [0-18112] | 6.7 (6.4)  5^c,d^ [0-35] | 89^c,d^ (93.7) | 907.6 (1021.3)  509.3^c^ [0-3247.7] | 4.6 (4.1)  4^c,d^ [0-13] | 13 (76.5) |
| Dural Nodule | Gd+ Delayed 3T FLAIR | 15.5 (30.2)  0^c^ [0-109] | 0.6 (0.9)  0^b,c^ [0-3] | 20^b,c^ (33.3) | 9.5 (17.9)  0 [0-67] | 0.4 (0.6)  0 [0-2] | 6 (37.5) |
|  | Gd+ Early 7T FLAIR | 32.2 (75.1)  6.2 [0-628.7] | 1.1 (1.4)  1^b,d^ [0-9] | 55^b^ (57.9) | 25.9 (56.6)  0 [0-226.5] | 0.9 (1.3)  0 [0-4] | 7 (41.2) |
|  | Gd+ Delayed 7T FLAIR | 28.6 (37.8)  12.6^c^ [0-207.9] | 1.9 (2.2)  1^c,d^ [0-8] | 67^c^ (70.5) | 16.6 (27.8)  1.7 [0-96.4] | 1 (1.2)  1 [0-4] | 9 (52.9) |
| Total | Gd+ Delayed 3T FLAIR | 445.1 (985.7)  109.7^b,c^ [0-109.7] | 3.2 (3.9)  2^b,c^ [0-19] | 42^b,c^ (70) | 358.1 (751.8)  22.9 [0-2889] | 2.8 (3.6)  1^c^ [0-12] | 11 (68.8) |
|  | Gd+ Early 7T FLAIR | 800.7 (1379.8)  249.3^b,d^ [0-9919.9] | 5.4 (4.6)  4^a,b,d^ [0-22] | 85^b,d^ (89.5) | 198.2 (252.2)  115.3 [0-866.4] | 2.7 (2.7)  1**^a,d^** [0-8] | 12 (70.6) |
|  | Gd+ Delayed 7T FLAIR | 1287.6 (2086.9)  646.7^c,d^ [0-18134.2] | 10.1 (6.9)  8^c,d^ [0-36] | 94^a,c,d^ (98.9) | 968.1 (1064.7)  605.7^c,d^ [0-3447.1] | 6.6 (4.9)  7^c,d^ [0-18] | 15^a^ (88.2) |

HC = healthy control, MS = multiple sclerosis, ME = meningeal enhancement

Values shown are mean (SD) and median [minimum-maximum]. Mean shown for informational purposes, but statistical tests performed using non-parametric testing, and thus statistical significance to be shown for median values.

“**^a^**” = p < 0.05 for comparison of HC and MS, “^b^” = p < 0.05 for comparison between Gd+ Delayed 3T FLAIR and Gd+ Early 7T FLAIR, “^c^” = p < 0.05 for comparison between Gd+ Delayed 3T FLAIR and Gd+ Delayed 7T FLAIR, “^d^” = p < 0.05 for comparison of Gd+ Early 7T FLAIR and Gd+ Delayed 7T FLAIR.

Table S2: Demographic and clinical characteristics by presence of LMPE in the MS cohort.

|  | | LMPE | | | | | |
| --- | --- | --- | --- | --- | --- | --- | --- |
|  | | Gd+ 3T  LMPE +  N = 38 | Gd+ 3T LMPE -  N = 22 | Gd+ Early 7T  LMPE +  N = 79 | Gd+ Early 7T LMPE -  N = 16 | Gd+ Delayed 7T  LMPE +  N = 94 | Gd+ Delayed 7T LMPE -  N = 1 |
| Mean Age (SD) | | 45.4 (10.4) | 44.5 (9.9) | 46.4* (10.5) | 39.6* (9.0) | 45.2 (10.6) | 53 (n/a) |
| Median Disease Duration [min-max] | | 14.0 [1.8-33.3] | 12.5 [0.4-24.2] | 11.2 [1.5-47.8] | 12.4 [0.44, 23.4] | 11.3 [0.4-47.8] | 17.9 [n/a] |
| Female Gender (%) | | 30* (79.0) | 11* (50) | 54 (68.4) | 12 (75) | 65 (69.1) | 1 (n/a) |
| Disability Scales | Median EDSS  [min-max] | 2.5 [0-6.5] | 2.25 [0-6.5] | 2.5 [0-6.5] | 2.5 [0-6.5] | 2.5 [0-6.5] | 4.0 [n/a] |
|  | Median 9HPT  [min-max] | 21.2 [15.6-425.7] | 22.4 [17.6-231.1] | 22.6 [15.6-425.7] | 22.6 [17.6-65.0] | 22.6 [15.6-425.7] | 20.5 [n/a] |
|  | Median T25W [mix-max] | 4.9 [3.7-161] | 5.0 [3.7-161] | 5.4 [3.2-161] | 5.1 [3.4-24.1] | 5.3 [3.2-161] | 6.4 [n/a] |
|  | Mean SDMT (SD) | 55.0 (16.0) | 52.2 (12.5) | 53.31 (15.05) | 57.55 (15.27) | 54.0 (15.1) | 49 (n/a) |
|  | Median PASAT [min-max] | 47 [0-59] | 50 [26-58] | 45 [0, 60] | 42.5 [23, 55] | 45 [0-60] | 49 [n/a] |

LMPE = leptomeningeal and paravascular enhancement, SD = standard deviation. EDSS = Expanded Disability Status Scale, 9HPT = Nine Hole Peg Test (in seconds), T25W = Timed 25-foot walk (in seconds), SDMT = Symbol Digit Modalities Test (# correct).

Wilcoxon rank sum test was used for all values shown as median, t-test for those shown as mean, and proportions were tested by Chi-square or Fisher’s exact test. * = p < 0.05. Note: statistical tests not completed for Gd+ Delayed 7T, as only 1 subject in LMPE- group. SD and range shown as n/a for the same reason.
